# RenalTransLSTM: Multi-Horizon Prediction of Acute Kidney Injury in ICU Patients using a Hybrid LSTM-Transformer Architecture

**DOI:** 10.64898/2026.07.02.26357177

**Authors:** S. M. Saiful Islam Badhon, Mohammad Adibuzzaman, Abu Saleh Mohammad Mosa, Serdar Bozdag, Ana D. Cleveland, Junhua Ding, K S M Tozammel Hossain

## Abstract

**Objective:** Acute kidney injury (AKI) affects a large proportion of patients in the intensive care unit (ICU) and is a major contributor to morbidity, mortality, and cost. Although electronic health records (EHRs) capture rich longitudinal data, many predictive models fail to detect AKI early enough for effective intervention. Non-temporal methods such as logistic regression and XGBoost treat patient history as aggregated risk factors, discarding the temporal evolution of clinical state. A recent trend is to employ temporal models, such as recurrent neural networks, to capture sequential patterns, but these models struggle with irregular sampling and limited long-range contextual awareness. To address the challenge, we propose RenalTransLSTM, a hybrid temporal deep learning framework for early, multi-horizon AKI prediction and identification of modifiable risk factors.

**Methods:** RenalTransLSTM integrates Long Short-Term Memory (LSTM) networks with Transformer encoders to model both local temporal dynamics and global contextual dependencies in ICU time-series data. Using 48-hour patient histories from MIMIC-IV (61,735 admissions), the model predicts AKI at 6-, 12-, and 24-hour lead times. We benchmark the model against SVM, XGBoost, LSTM, TG-LSTM, and a Transformer, and apply Integrated Gradients and counterfactual analysis to identify modifiable risk factors.

**Results:** RenalTransLSTM outperforms all baselines across most horizons and metrics, achieving AUROC above 0.90 and F1-scores reaching 0.85 while maintaining balanced precision and recall on imbalanced data. Ablation studies confirm that combining LSTM and Transformer components improved robustness and predictive performance. Counterfactual analysis identifies clinically meaningful, modifiable risk factors associated with AKI progression.

**Conclusion:** RenalTransLSTM offers an effective, interpretable framework for early AKI prediction in the ICU, supporting proactive intervention and clinical decision support.

## 1. Introduction

Acute kidney injury (AKI) is a clinical syndrome characterized by rapid deterioration of kidney function occurring over hours to days. The syndrome affects approximately 13.3 million people annually worldwide, with nearly 85% of cases occurring in low- and middle-income countries [18, 9]. In high-income countries, AKI incidence ranges from 3,000 to 5,000 cases per million population annually [9]. The burden is particularly severe in intensive care units (ICUs), where more than 50% of patients develop AKI [7]. As there is no effective treatment for AKI [26, 1, 28, 10], early prevention through timely identification of at-risk patients remains the most viable clinical strategy. Early intervention can avert kidney failure, reduce the need for renal replacement therapy, improve patient outcomes, and alleviate the substantial burden on healthcare systems [10].

The widespread adoption of electronic health records (EHRs) in hospital systems has created new opportunities to leverage machine learning for early disease prediction, including AKI [31]. However, most existing AKI prediction models are non-temporal, such as logistic regression, support vector machines (SVMs), and gradient-boosting methods [23]. These approaches treat the patient’s history as a collection of aggregated risk factors, ignoring the sequential evolution of clinical states. Although effective for retrospective analysis, they fail to capture critical temporal dependencies for prospective prediction. Recent efforts have introduced temporal models, including recurrent neural networks (RNNs) and long short-term memory (LSTM) networks, to learn sequential patterns from EHR time series [31, 19, 3, 17, 11, 37]. Yet these models face two fundamental limitations: difficulty in maintaining long-range contextual information and sensitivity to the irregular sampling characteristic of ICU data. Moreover, many temporal approaches lack a clearly defined buffer between the observation window and the prediction target (Figure 1), potentially generating forecasts with insufficient lead time for clinical action.

**Figure 1:**
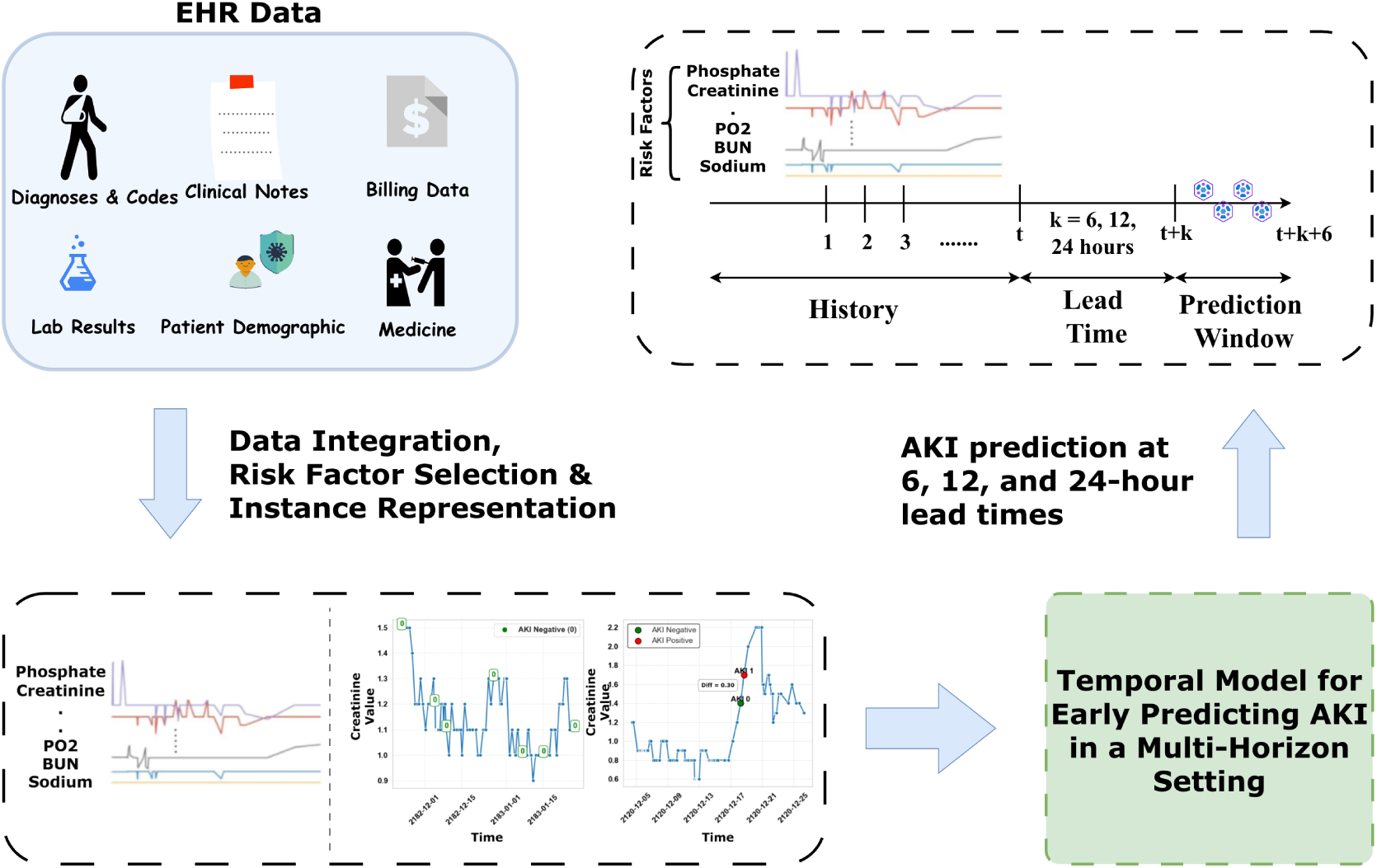
A framework for generating early warnings for AKI in the ICU using EHR streams. This pipeline enables proactive AKI prediction before clinical onset. Heterogeneous EHR data are integrated, temporally structured, and transformed into fixed windows that feed a hybrid LSTM–Transformer model to anticipate AKI at 6, 12, and 24 hours in advance.

To address these limitations, we propose *RenalTransLSTM*, a hybrid deep learning framework that integrates LSTM networks with Transformer encoders. The LSTM component captures local temporal dynamics and sequential dependencies, while the Transformer’s self-attention mechanism models global contextual relationships across the patient history. By combining these architectures, RenalTransLSTM overcomes the individual weaknesses of each approach. The framework incorporates explicit lead times of 6, 12, and 24 hours between the observation window and the prediction target, ensuring that forecasts provide clinicians with actionable time for preventive intervention. Figure 1 illustrates the overall pipeline. The key contributions of this study are:

- **A hybrid temporal architecture for AKI forecasting with lead time:** We develop RenalTransLSTM, which augments LSTM-based sequence learning with Transformer encoders to capture both local temporal dynamics and global feature dependencies in EHR time series, enabling more accurate early detection of AKI.
- **Clinically actionable prediction horizons:** The framework generates AKI forecasts at 6-, 12-, and 24-hour lead times, providing clinicians with sufficient time to implement preventive measures.
- **Comprehensive empirical validation:** We evaluate RenalTransLSTM on the MIMIC-IV dataset comprising approximately 60,000 ICU admissions, demonstrating superior performance over baseline methods through extensive experiments and ablation studies that confirm the contribution of each architectural component.
- **Modifiable risk factors:** We validate our model’s interpretability using two distinct methods. Both of these methods identify clinically meaningful and modifiable risk factors as important predictors.

### Statement of Significance

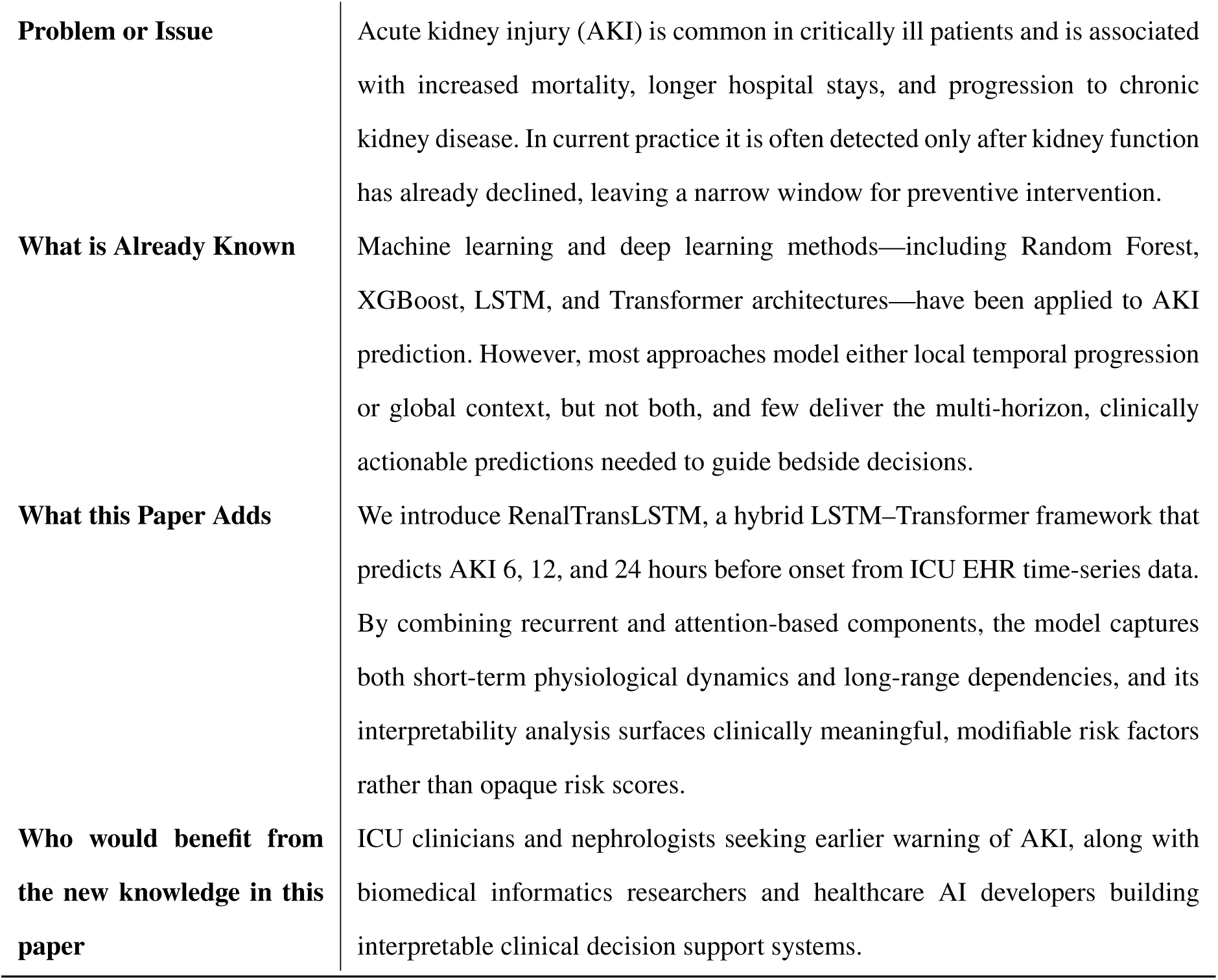

|  |  |
| --- | --- |
| **Problem or Issue** | Acute kidney injury (AKI) is common in critically ill patients and is associated with increased mortality, longer hospital stays, and progression to chronic kidney disease. In current practice it is often detected only after kidney function has already declined, leaving a narrow window for preventive intervention. |
| **What is Already Known** | Machine learning and deep learning methods—including Random Forest, XGBoost, LSTM, and Transformer architectures—have been applied to AKI prediction. However, most approaches model either local temporal progression or global context, but not both, and few deliver the multi-horizon, clinically actionable predictions needed to guide bedside decisions. |
| **What this Paper Adds** | We introduce RenalTransLSTM, a hybrid LSTM–Transformer framework that predicts AKI 6, 12, and 24 hours before onset from ICU EHR time-series data. By combining recurrent and attention-based components, the model captures both short-term physiological dynamics and long-range dependencies, and its interpretability analysis surfaces clinically meaningful, modifiable risk factors rather than opaque risk scores. |
| **Who would benefit from the new knowledge in this paper** | ICU clinicians and nephrologists seeking earlier warning of AKI, along with biomedical informatics researchers and healthcare AI developers building interpretable clinical decision support systems. |

## 2. Literature Review

### 2.1. Traditional Machine Learning Approaches for AKI Prediction

Traditional machine learning (ML) approaches have been widely employed to predict AKI across diverse patient populations and clinical scenarios. These methods typically rely on feature selection techniques and engineered predictors to build interpretable models. Here we review key studies using traditional ML algorithms for AKI prediction, emphasizing their datasets, methodologies, and outcomes. Lin et al. (2024) utilized the MIMIC-III database to develop ML models for predicting AKI in critically ill septic patients. Feature selection was performed using the Boruta algorithm, and models such as logistic regression (LR), k-nearest neighbors (KNN), support vector machines (SVM), decision trees, random forests (RF), artificial neural networks (ANN), and extreme gradient boosting (XGBoost) were constructed using tenfold cross-validation [19]. Among the models, XGBoost achieved the highest AUC (0.821), outperforming traditional clinical scores like SOFA and SAPS II. The study demonstrated the efficacy of ML in identifying high-risk patients early and providing actionable insights for intervention. Cai et al. (2022) employed ML techniques to predict AKI in patients with acute myocardial infarction (AMI) using the MIMIC-III and MIMIC-IV databases. Models such as RF, Naive Bayes (NB), SVM, XGBoost, decision trees (DT), and LR were constructed [3]. Using SHapley Additive exPlanations (SHAP), serum creatinine, partial prothrombin time, and blood glucose were identified as key predictors. RF demonstrated the highest AUC (0.781) on external validation, underscoring its effectiveness in risk stratification. Kim et al. (2022) developed a risk scoring system for vancomycin-associated AKI based on logistic regression, elastic net, RF, and SVM [17]. Using medical records of 104 patients undergoing vancomycin therapy, the study incorporated nephrotoxic agents and vancomycin trough levels as key predictors. SVM achieved an AUC of 0.739, highlighting its adaptability to clinical scenarios involving nephrotoxic drugs. The scoring system enabled clinicians to estimate AKI risk in real-time, facilitating personalized treatment adjustments. Hsu et al. (2022) constructed an SVM model for predicting AKI risk in ultramarathon runners based on prerace data, including blood and urine tests. The SVM model achieved an accuracy of 90% in cross-validation, with high sensitivity (90%) and specificity (100%) [11]. This study showcased the potential of ML in niche populations, where tailored predictors such as muscle mass and hydration status are crucial. Yang et al. (2022) developed ML models for predicting AKI in patients with severe acute pancreatitis (SAP) using data from 424 patients [37]. Models including RF, XGBoost, ANN, and DT were evaluated, with RF achieving the highest AUC (0.902) by incorporating inflammatory markers such as C-reactive protein (CRP) and neutrophil-to-albumin ratio (NAR). This study highlighted the importance of integrating inflammatory factors into AKI prediction models. Omar et al. (2024) analyzed ML models for predicting AKI necessitating dialysis after cardiac surgery using electronic health records from a Malaysian center [23]. Gradient boosted trees (GBT) demonstrated the best performance, achieving an AUC of 94.61% and sensitivity of 91.30%. RF also performed well, with an AUC of 94.78%, while SVM achieved higher sensitivity (98.57%) but lower specificity (59.55%). These findings illustrate the need for model selection based on specific clinical goals. Qian et al. (2021) used MIMIC-III data to compare six ML models, including LR, SVM, RF, XGBoost, LightGBM, and convolutional neural networks (CNN). LightGBM achieved the highest AUC (0.905) and recall (0.836), followed by XGBoost. The study emphasized the superiority of ensemble methods for early AKI detection, particularly in leveraging serum creatinine as the most significant predictor.

Above works on AKI prediction rely on traditional machine learning models such as Random Forest, XGBoost, and SVM, which primarily operate in a non-temporal setting. These models focus on static feature selection or aggregate patient data, limiting their ability to capture dynamic trends and sequential dependencies inherent in ICU records. Our proposed hybrid LSTM-Transformer model addresses these limitations by leveraging time-series data to model both local temporal patterns and global contextual relationships, enabling more comprehensive and clinically relevant predictions.

### 2.2. Time-Aware and Deep Learning Approaches for AKI and Related Predictions

The following section explores the emergence of time-aware and deep learning approaches that address these limitations by leveraging sequential and time-series data. These advanced techniques, such as recurrent neural networks (RNNs), long short-term memory (LSTM) networks, and transformers, provide a more dynamic modeling framework for capturing patient trajectories over time. These models go beyond the capabilities of traditional ML approaches by directly incorporating time-dependent factors and offering the potential for multi-horizon predictions, including AKI prediction at various time points before its occurrence. As we delve into these modern methodologies, we will examine how they build upon the foundations laid by traditional machine learning. Below, we summarize key studies that highlight the role of these advanced techniques in healthcare predictions, including AKI and related conditions.

Zhang et al. (2021) utilized EHR audit log data to predict next-day hospital discharge [38]. By modeling granular interactions between users and EHR systems, their deep learning model achieved an AUROC of 0.921, demonstrating the potential of interaction data for outcome predictions. However, their model focuses on non-physiological data, limiting its applicability to AKI and similar conditions. Zeng et al. (2022) proposed a time-aware attention model for predicting AKI after pediatric cardiac surgery using sequential time-series clinical data. Their model, evaluated on the PIC database, achieved an AUROC of 0.908, outperforming traditional methods. While effective, their approach was tailored to pediatric populations and lacks generalizability across other clinical settings. Zisser and Aran (2023) introduced STRAFE, a Transformer-based survival analysis model for predicting chronic kidney disease (CKD) deterioration [39]. Using a large claims dataset, STRAFE demonstrated superior performance for time-to-event predictions, outperforming fixed-time models. Despite its effectiveness, STRAFE is limited to survival analysis and does not address multi-horizon predictions. Bashiri et al. (2023) evaluated deep learning models (LSTM/GRU, temporal convolutional networks, and CNNs) for variable-length time-series tasks, including AKI prediction [2]. Their results showed LSTM/GRU with piece-wise linear encoding achieving the highest AUROC (0.92 for AKI). However, their reliance on pre-defined encoding strategies may limit adaptability to diverse ICU datasets. Catling and Wolff (2019) utilized TCNs to predict ICU events within 1–6 hours, achieving significant improvements in sensitivity and positive predictive value [5]. While TCNs effectively handle sequential data, their capacity to capture long-term dependencies is limited compared to Transformer-based models. Roy et al. (2021) introduced SeqSNR, a multitask learning framework using MIMIC-III data to predict ICU outcomes, including AKI and mortality [25]. SeqSNR achieved modest gains in AUROC compared to single-task models, but its computational complexity may hinder real-time clinical applications. Huang et al. (2021) proposed Deep Significance Clustering (DICE) to identify risk-stratified subgroups in heterogeneous populations, including AKI patients [13]. DICE demonstrated high interpretability and predictive accuracy but was designed for clustering rather than direct time-series predictions. Olaimat and Bozdag (2023) developed TA-RNN for handling irregular time intervals in EHR data, incorporating time embeddings and attention mechanisms [22]. Tested on MIMIC-III, TA-RNN achieved superior sensitivity and interpretability, highlighting influential visits and features. However, its primary application is in mortality prediction, not AKI.

These studies illustrate the potential of deep learning and time-aware methods for clinical predictions. However, limitations persist. Many approaches, such as TA-RNN and SeqSNR, focus on non-specific or single-task outcomes rather than multi-horizon AKI predictions. Time-to-event models like STRAFE lack the flexibility to predict specific horizons (e.g., 6, 12, or 24 hours). Few studies integrate both local temporal trends and global contextual relationships in ICU time-series data. Our proposed hybrid LSTM-Transformer architecture combines the strengths of sequential modeling (LSTM) and global context awareness (Transformer). Unlike existing methods, it explicitly targets multi-horizon AKI prediction (6, 12, and 24 hours) and adapts to the dynamic nature of ICU time-series data. By capturing both short-term dependencies and long-term relationships, our model provides interpretable and clinically actionable insights, addressing key gaps in the current literature.

## 3. Methods

### 3.1. Problem Formulation

We develop a temporal model to anticipate AKI at least six hours before onset, using the most recent 48 hours of patient history. The model exploits sequential clinical measurements and demographic data to predict AKI risk. Each patient history is represented as a sequential time-series observations of physiological and clinical risk factors collected over the last 48 hours. Each time step includes measurements of several risk factors, including creatinine levels, blood urea nitrogen (BUN), temperature, pH levels, potassium, plateau pressure, hemoglobin etc.

We cast the AKI risk anticipation as a binary classification problem, where the outcome “yes” indicates that the patient is likely to develop AKI after a predefined lead time (e.g., 6, 12, or 24 hours) and the outcome “no” indicates that AKI is unlikely to occur after the lead time.

Let *X* = {*x*_1_, *x*_2_, …, *x_T_* } be a sequence of patient data over *T* time steps (e.g., 48 hours with each hour as a time step). Here, *X_t_* ∈ ℝ*^d^* represents the vector of risk factors at time *t*, where *d* is the number of features (e.g., clinical measurements and demographic information). Let *Y_t_*_+*k*_ ∈ {0, 1} be the target variable for AKI prediction after the specified lead time window *k* (e.g., *k* = 6, 12, 24 hours).

The objective is to learn a function:

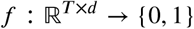

that maps the input time series *X* to the binary target *y*, maximizing predictive accuracy while balancing sensitivity (recall) and specificity (precision).

### 3.2. Baselines

Several models could be considered as baselines for this problem. These models are broadly categorized into two groups: non-temporal and temporal.

#### 3.2.1. Non-Temporal Methods

Non-temporal methods treat the patient history as independent observations, ignoring the sequential dependencies between risk factors. Although discarding time information is a limitation, this type of method offers better explainability. We consider support vector machine (SVM) and gradient boosting in this category.

##### Support Vector Machine (SVM)

SVM is a supervised learning method that seeks to identify the optimal hyperplane separating the two classes AKI and non-AKI. We use all the independent risk factors. One of the key parameters of SVM is kernel, for which we use the Radial Basis Function (RBF) to capture non-linear relationships. The reason for choosing SVM is that it handles complex decision boundaries, especially in high-dimensional settings [27].

##### Gradient Boosting (GB)

Gradient Boosting is an ensemble learning algorithm that builds a predictive model by combining multiple weak learners (decision trees). We used the same risk factors as SVM. The number of estimators, learning rate, and maximum tree depth were optimized using grid search. GB is a good candidate for a classification problem with highly imbalance data [35].

#### 3.2.2. Temporal Methods

Temporal methods exploit the sequential nature of time-series data, capturing rich patterns and dependencies over time. These methods are inherently better suited for AKI prediction, as they can analyze trends and changes in risk factors.

##### Feedforward Neural Network

Feedforward neural networks (FNNs), also known as multilayer perceptrons (MLPs), consist of fully connected layers that process input data without any notion of sequence order. When applied to time-series data, the temporal dimension is flattened into a single feature vector, allowing the network to learn nonlinear combinations of features across all time steps. FNNs are computationally efficient and serve as a useful baseline for assessing whether temporal structure contributes meaningfully to predictive performance. However, by discarding the sequential ordering of measurements, FNNs are unable to model how clinical variables evolve over time, limiting their ability to capture trends such as the progressive rise in creatinine levels preceding AKI [12].

##### Recurrent Neural Network (RNN)

Recurrent neural networks (RNNs) process sequential data by maintaining a hidden state that is updated at each time step, allowing information from earlier time points to influence later predictions. This architecture enables RNNs to capture short-term temporal dependencies and sequential patterns in clinical time-series data. However, standard RNNs are prone to vanishing and exploding gradient problems during training, which restricts their capacity to retain information over longer sequences. As a result, RNNs often struggle to capture long-range dependencies, motivating the development of gated architectures such as LSTM [4].

##### TG-LSTM

TG-LSTM introduces an additional temporal gating mechanism to dynamically weigh the significance of each time step in the sequence. TG-LSTM improves temporal modeling by accounting for irregular patterns, such as sudden spikes in BUN or hemoglobin levels. While TG-LSTM enhances time-step relevance, it lacks the ability to capture broader contextual relationships like Transformers [12].

### 3.3. Proposed Methods

We develop a hybrid LSTM-Transformer model for predicting Acute Kidney Injury (AKI) in ICU patients. The proposed model leverages the strengths of both architectures, combining the sequential learning capabilities of LSTM with the contextual analysis of the Transformer. Our motivation for this approach comes from works such as Hybrid LSTM-Transformer in Engineering Systems (engineering systems) [4], Clinical Time Series Analysis using Attention Models (clinical time-series data) [29], and TransLSTM: A hybrid LSTM-Transformer model for fine-grained suggestion mining (suggestion mining) [24], which demonstrate the benefits of integrating LSTM and Transformer for improved accuracy and interpretability.

#### Transformer Encoder Layers

The Transformer encoder applies multi-head self-attention to capture relationships between all time steps in the input sequence:

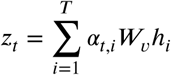

where

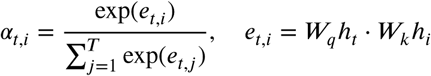

*W_q_*, *W_k_*, *W_v_* are learnable projection matrices for queries, keys, and values. Residual connections and layer normalization are applied to ensure stability:

Attention Output = LayerNorm(Dropout(MultiHeadAttention)+Input)

Feed Forward Output = LayerNorm(Dropout(Dense)+ Attention Output)

*LSTM Component:* The LSTM processes the input sequence to learn temporal dependencies:

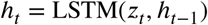

where ℎ*_t_* ∈ ℝ*^m^* is the hidden state at time *t*, and *m* is the dimensionality of the hidden state. This component effectively captures trends, such as gradual increases in creatinine levels or consistent changes in BUN values.(see Fig. 3b)

## 4. Results

We design experiments to address the following research questions:

- **RQ1.** How do the proposed methods fare against the baselines? (see Sec. 4.4)
- **RQ2.** Are the proposed methods robust in terms of parameter variation? (see Sec. 4.5)
- **RQ3.** How do various components of the method contribute to the performance? (see Sec. 4.6)
- **RQ4.** Does the proposed model identify clinically meaningful modifiable risk factors? (see Sec. 4.7)

### 4.1. Datasets

We use an extensive, openly accessible MIMIC-IV EHR dataset to train and evaluate our models. The Medical Information Mart for Intensive Care IV (MIMIC-IV version 1.0) database was developed at the Massachusetts Institute of Technology (MIT) and Beth Israel Deaconess Medical Center. This dataset includes rich information about patients with their consent. Data collection was passive, with no impact on patient safety, and all information was de-identified in accordance with HIPAA’s Privacy Rule. Since research involving de-identified data is classified as nonhuman subject research, this study did not require additional institutional or ethical approvals [8]. The timeline of this dataset ranges from 2008 to 2019. This dataset includes data on both the ICU and the outpatient settings. This study considers ICU patients with detailed records of 61,735 admissions from a single hospital.

#### Cohort Selection

The cohort selection process began by including all adult admissions (patients aged ≥18), resulting in an initial cohort of 61,735 admissions. This age-based filtering standardizes the cohort to adults, as physiological and clinical characteristics can vary significantly between adults and minors, impacting the generalizability of findings [20]. Admissions with at least one recorded observation for each selected risk factor are retained, reducing the cohort to 21,100. Ensuring each admission includes data on all relevant risk factors helps mitigate missing-data issues, improving model reliability and reducing bias [21]. We then select admissions with a continuous observation window of 60 hours, using the first 48 hours of history for model training, yielding a final cohort of 18,131 admissions. This consistent time frame enables the model to capture relevant temporal patterns and early indicators of risk, as variable observation windows could introduce noise and reduce predictive accuracy [15]. Such structured cohort selection ensures a robust dataset, enhancing both the interpretability and performance of predictive models, and aligns with best practices in healthcare data modeling [21].

We annotate each history as AKI positive (i.e., presence of AKI) or AKI negative (i.e., absence of AKI) using the KDIGO formula [32] (see Fig. 3a).

### 4.2. Risk Factor Selection

We extract numerical risk factors from ICU charts (6,463 features) and lab events (726 features). Using the KDIGO [32] criteria (increase in serum creatinine by ≥ 0.3 mg/dl within 48 hours), we label AKI (0/1) using patient’s creatinine and chart time values. Given this labeled data, we estimate pairwise Pearson’s correlation coefficients between the AKI labels and each feature. Finally, we select 52 statistically significant features.

When we exclude serum creatinine, the model achieved moderate performance (F1 score: 0.71, Accuracy: 0.78), highlighting the significant role of other risk factors. However, including SC alongside other variables led to a substantial improvement (F1 score: 0.84; accuracy: 0.88). Further analysis of SC’s contribution, using feature importance or ablation studies, could help optimize its role in prediction.

### 4.3. Experimental Setup

We evaluate both non-temporal and temporal models for kidney injury prediction. For non-temporal models, we aggregate patient history and create a vector of risk factors. We use 80% of the data for training and validation and 20% for unbiased testing. We apply 5-fold cross-validation and select the best model for unbiased evaluation.

### 4.4. Identify the Predictive Power of the Proposed Method (RQ1)

We train and evaluate our proposed method and compare it with existing methods (see Table 2). The proposed method outperforms other methods across most performance measures. The Trans-LSTM demonstrates improved recall, indicating its ability to correctly identify more AKI instances. Overall, the method performs better in all the scenarios across the three time windows (6 hours, 12 hours, and 24 hours) and four evaluation metrics (precision, recall, F1-score, accuracy, ROC, and PRC), highlighting its effectiveness in handling both precision and recall trade-offs in imbalanced data. The Transformer model outperforms the proposed Transformer LSTM in precision. This phenomenon is due to the dataset’s imbalance, with non-AKI instances significantly outnumbering AKI instances. This imbalance allows the Transformer to better capture non-AKI cases, resulting in fewer false positives (FPs).

**Table 1.** Feature Characteristics. This table compares the patient characteristics between the AKI and Non-AKI groups. Additionally, the P-values for each feature are calculated to assess their relevance in predicting AKI.

| Features | Overall<br>*Median (Min,Max) or Count* | Non-AKI<br>*Median (Min,Max) or Count* | AKI<br>*Median (Min,Max) or Count* | P-Value |
| --- | --- | --- | --- | --- |
| Gender | 18131 | 11306 | 6825 | 5.72E-10 |
| M | 10110 | 6103 | 4007 |  |
| F | 8021 | 5203 | 2818 |  |
| Admission Type | 18131 | 11306 | 6825 | 2.60E-59 |
| Direct Emer | 718 | 339 | 379 |  |
| Direct Observation | 2 | 2 | 0 |  |
| Elective | 497 | 252 | 245 |  |
| EU Observation | 1 | 0 | 1 |  |
| EW Emer | 9689 | 6479 | 3210 |  |
| Observation Admit | 2331 | 1490 | 841 |  |
| Surgical Same Day Admission | 1245 | 629 | 616 |  |
| Urgent | 3648 | 2115 | 1533 |  |
| Age | 65.0 (18, 91) | 66.0 (18, 91) | 65.0 (18, 91) | 9.80E-11 |
| Plateau Pressure | 19.0 (0.0, 46.0) | 19.0 (0.0, 46.0) | 19.0 (0.0, 42.0) | 2.29E-13 |
| BUN | 22.0 (1.0, 160.0) | 22.0 (2.0, 149.0) | 22.0 (1.0, 160.0) | 5.29E-90 |
| PTT | 33.7 (17.6, 150.0) | 33.7 (17.8, 150.0) | 33.7 (17.6, 150.0) | 1.05E-05 |
| Phosphorous | 3.5 (0.6, 12.9) | 3.5 (1.0, 12.9) | 3.5 (0.6, 10.5) | 6.01E-71 |
| Dialysis patient | 0.0 (0.0, 1.0) | 0.0 (0.0, 1.0) | 0.0 (0.0, 0.0) | >0.0002 |
| Goal Richmond-RAS Scale | 0.0 (0.0, 5.0) | 0.0 (0.0, 5.0) | 0.0 (0.0, 5.0) | <0.002 |
| CAM-ICU MS Change | 0.0 (0.0, 1.0) | 0.0 (0.0, 1.0) | 0.0 (0.0, 1.0) | 0.08 |
| Alveolar-arterial Gradient | 501.0 (372.0, 635.0) | 501.0 (372.0, 635.0) | 501.0 (394.0, 606.0) | 0.86 |
| Hemoglobin | 9.8 (4.1, 21.1) | 9.8 (5.6, 16.1) | 9.8 (4.1, 21.1) | <0.002 |
| Lactate | 2.0 (0.4, 16.4) | 2.0 (0.5, 16.4) | 2.0 (0.4, 15.5) | <0.002 |
| pCO2 | 40.0 (15.0,107.0) | 40.0 (15.0,100.0) | 40.0 (18.0,107.0) | <0.002 |
| pH | 7.36 (1.0,9.0) | 7.36 (1.0,7.62) | 7.36 (3.0,9.0) | >0.002 |
| pO2 | 104.0 (21.0,508.0) | 104.0 (21.0,508.0) | 104.0 (21.0,457.0) | 2.09E-06 |
| Potassium, Whole Blood | 4.2 (2.8,7.3) | 4.2 (2.8,7.3) | 4.2 (3.0,5.9) | 0.39 |
| Required O2 | 84.0 (67.0,100.0) | 84.0 (67.0,100.0) | 84.0 (69.0,99.0) | 0.79 |
| Temperature | 36.9 (32.8,40.0) | 36.9 (32.8,40.0) | 36.9 (34.0,39.4) | 0.27 |
| Anion Gap | 14.0 (3.0,35.0) | 14.0 (3.0,35.0) | 14.0 (5.0,29.0) | 5.24E-69 |
| Bilirubin, Direct | 1.8 (0.1,22.5) | 1.8 (0.1,22.5) | 1.8 (0.1,5.8) | 0.36 |
| Bilirubin, Indirect | 1.0 (0.1,12.0) | 1.0 (0.1,12.0) | 1.0 (0.1,8.1) | 0.77 |
| Bilirubin, Total | 0.9 (0.0,54.0) | 0.9 (0.0,51.6) | 0.9 (0.1,54.0) | >0.001 |
| Calcium, Total | 8.2 (0.0,13.7) | 8.2 (0.0,13.7) | 8.2 (4.9,12.6) | 5.26E-05 |
| Cholesterol, HDL | 43.0 (8.0,74.0) | 43.0 (26.0,67.0) | 43.0 (8.0,74.0) | 0.09 |
| Cholesterol, LDL, Calculated | 76.0 (27.0,156.0) | 76.0 (27.0,118.0) | 76.0 (33.0,156.0) | 0.52 |
| Cholesterol, Total | 148.0 (67.0,397.0) | 148.0 (67.0,181.0) | 148.0 (76.0,397.0) | 0.99 |
| Creatinine | 1.0 (0.0,20.7) | 1.8 (0.2,20.7) | 0.8 (0.0,18.0) | 0 |
| Iron Binding Capacity, Total | 209.0 (52.0,417.0) | 209.0 (116.0,404.0) | 209.0 (52.0,417.0) | 0.53 |
| Lactate Dehydrogenase (LD) | 318.0 (65.0,12845.0) | 318.0 (118.0,12845.0) | 318.0 (65.0,7460.0) | 1.37E-05 |
| Magnesium | 2.0 (0.2,4.7) | 2.0 (0.2,4.5) | 2.0 (0.8,4.7) | 2.12E-22 |
| Phosphate | 3.4 (0.6,15.3) | 3.4 (0.9,15.3) | 3.4 (0.6,10.9) | 6.15E-117 |
| Potassium | 4.1 (1.8,8.6) | 4.1 (1.8,6.6) | 4.1 (2.1,8.6) | 6.57E-49 |
| Sodium | 138.0 (115.0,170.0) | 138.0 (115.0,170.0) | 138.0 (116.0,162.0) | 1.21E-13 |
| Transferrin | 161.0 (40.0,321.0) | 161.0 (89.0,311.0) | 161.0 (40.0,321.0) | 0.53 |
| Urea Nitrogen | 22.0 (1.0,180.0) | 22.0 (2.0,169.0) | 22.0 (1.0,180.0) | 3.63E-155 |
| Vancomycin | 15.7 (2.8,47.8) | 15.7 (3.5,47.8) | 15.7 (2.8,37.0) | 7.03E-05 |
| Chloride, Urine | 45.0 (10.0,213.0) | 45.0 (10.0,160.0) | 45.0 (10.0,213.0) | 0.50 |
| Osmolality, Urine | 377.0 (55.0,899.0) | 377.0 (55.0,753.0) | 377.0 (64.0,899) | 0.50 |
| Eosinophils | 0.32 (0.0, 21.4) | 0.32 (0.0, 21.4) | 0.32 (0.0, 20.0) | 0.74 |
| INR(PT) | 1.3 (0.6, 9.2) | 1.3 (0.9, 9.2) | 1.3 (0.6, 7.6) | 2.92E-12 |
| MCV | 91.0 (55.0, 127.0) | 91.0 (57.0, 127.0) | 91.0 (55.0, 122.0) | >0.001 |
| Metamyelocytes | 0.0 (0.0, 20.0) | 0.0 (0.0, 20.0) | 0.0 (0.0, 8.0) | 0.729843 |
| Myelocytes | 0.0 (0.0, 14.0) | 0.0 (0.0, 13.0) | 0.0 (0.0, 14.0) | 0.948624 |
| Neutrophils | 81.1 (0.0, 97.1) | 81.1 (0.0, 97.0) | 81.1 (0.0, 97.1) | 0.056566 |
| PT | 14.6 (7.6, 95.9) | 14.6 (9.5, 95.9) | 14.6 (7.6, 79.3) | 4.47E-12 |
| RDW | 15.1 (11.1, 29.9) | 15.1 (11.6, 29.9) | 15.1 (11.1, 28.9) | 6.30E-26 |
| Glucose | 130.0 (28.0, 1000.0) | 130.0 (46.0, 657.0) | 130.0 (28.0, 1000.0) | 5.13E-12 |
| Ketone | 10.0 (10.0, 150.0) | 10.0 (10.0, 80.0) | 10.0 (10.0, 150.0) | 9.06E-05 |
| hours_since_icu_admit | 45.69 (1.14, 48.0) | 46.13 (2, 48.0) | 45.33 (2, 48.0) | 9.97E-85 |

**Table 2.** The proposed RenalTransLSTM consistently outperforms competing methods across most lead times and evaluation metrics. A = Accuracy; F1 = F1 Score; P = Precision; R = Recall; ROC = Receiver Operating Characteristic; PRC = Precision Recall Curve. Best results are highlighted in bold.

| Model | 6 Hours |  |  |  |  |  | 12 Hours |  |  |  |  |  | 24 Hours |  |  |  |  |  |
| --- | --- | --- | --- | --- | --- | --- | --- | --- | --- | --- | --- | --- | --- | --- | --- | --- | --- | --- |
|  | A | F1 | P | R | ROC | PRC | A | F1 | P | R | ROC | PRC | A | F1 | P | R | ROC | PRC |
| XG Boosting | .80 | .75 | .76 | .76 | .83 | .83 | .80 | .75 | .74 | .75 | .88 | .82 | .70 | .69 | .69 | .72 | .85 | .78 |
| SVM | .77 | .69 | .78 | .62 | .86 | .78 | .77 | .67 | .77 | .60 | .88 | .77 | .70 | .56 | .73 | .52 | .83 | .72 |
| Simple Feed Forward | .79 | .74 | .78 | .70 | .78 | .80 | .78 | .72 | .73 | .72 | .77 | .78 | .79 | .71 | .71 | .70 | .77 | .76 |
| RNN | .82 | .78 | .81 | .75 | .81 | .83 | .86 | .82 | .81 | .83 | .85 | .86 | .82 | .75 | .74 | .77 | .81 | .80 |
| TG LSTM | .79 | .76 | .73 | .79 | .79 | .80 | .78 | .75 | .75 | .75 | .85 | .81 | .73 | .72 | .70 | .75 | .86 | .79 |
| RenalTransLSTM | **.88** | **.85** | **.87** | **.84** | **.94** | **.93** | **.88** | **.84** | **.87** | **.80** | **.93** | **.92** | **.85** | **.81** | **.77** | **.84** | **.91** | **.90** |

The training loss consistently minimizes over time, while the validation loss remains close, indicating that the model improves across epochs and generalizes well to unseen data. Similarly, both training and validation accuracy follow parallel trends, demonstrating steady improvement. The minimal difference between the training and validation metrics, in both loss and accuracy, strongly suggests that the model is neither overfitting nor underfitting. This close alignment indicates that the model effectively learns the underlying patterns from the data and generalizes well to new, unseen instances, confirming robust model performance.(see Fig. 3c)

The model shows excellent performance in predicting the early onset of Acute Kidney Injury (AKI), with an AUC greater than 90%. This high AUC indicates that the model can effectively distinguish between patients who will develop AKI and those who will not, across various thresholds. Moreover, the PRC further reinforces the model’s capability, especially in imbalanced data, where negative cases (non-AKI) outnumber positive ones. With an Average Precision (AP) score above 90%, the model demonstrates high precision, ensuring that predicted AKI cases are truly at risk, and high recall, capturing most of the actual AKI cases. These metrics together highlight the model’s robustness in early AKI detection, making it a reliable tool for timely intervention and better patient outcomes. (see Fig. 3d)

### 4.5. Determine Robustness of the Proposed Method (RQ2)

We vary different parameters to assess the robustness of the proposed approach. The key parameters include learning rate and batch size.

#### 4.5.1. Learning Rate Variation

To evaluate the robustness of our proposed hybrid Transformer-LSTM model, we conduct experiments using three different learning rates (0.001, 0.0001, and 0.00001) across prediction horizons of 6, 12, and 24 hours. The results show that the learning rate 0.001 consistently delivered balanced precision, recall, and F1 scores across all horizons, making it the most robust setting. For the 6-hour horizon, this learning rate achieves an F1-score of 0.86; for the 12-hour and 24-hour horizons, it maintains strong performance with F1-scores of 0.84 and 0.80, respectively. Lower learning rates, such as 0.0001, yield comparable results for longer horizons but show lower recall for shorter predictions, whereas 0.00001 underperforms overall due to slower convergence and diminished precision.

#### 4.5.2. Batch Size Variation

We examine the effect of batch size (32, 64, and 128) across the same prediction horizons (6, 12, and 24 hours) to assess the model’s robustness and stability. A batch size of 64 provides the most balanced performance across all time horizons. For the 6-hour prediction horizon, the model achieves an F1-score of 0.84 with a batch size of 64, consistent with the performance observed at a learning rate of 0.001. For the 12- and 24-hour horizons, batch size 64 maintains strong F1 Scores of 0.84 and 0.80, respectively, consistent with the trends observed with the optimal learning rate.

In contrast, smaller batch sizes (32) yield slightly higher precision but lower recall, especially for longer prediction horizons. Larger batch sizes (128) show diminished precision but with higher recall, particularly at the 6-hour horizon. However, performance declines slightly at the 12- and 24-hour horizons with a batch size of 128, highlighting that larger batch sizes can sometimes trade off precision for recall.

### 4.6. Effect of Model Components (RQ3)

To assess the contribution of each component of the proposed model, we conduct an ablation study. An ablation study evaluates the impact of removing or altering specific parts of a model to determine their influence on overall performance. In our architecture, three key components are examined: (a) the Transformer, (b) the LSTM layer, and (c) the fully connected layers. The findings are summarized below.

#### 4.6.1. Removing the Transformer

When the Transformer module is removed, the model reduces to a standalone LSTM architecture (see Fig. 2 - B). The results are reported in Table 4. Without the Transformer, precision decreases across all prediction horizons, with the largest drop observed for the 24-hour prediction (from 0.81 to 0.64). Although recall remains relatively high for the 6-hour prediction (0.89), it declines substantially for longer horizons. This indicates that an LSTM-only model struggles to maintain balanced performance, especially at extended lead times, underscoring the importance of Transformer-based representations in capturing complex temporal dependencies.

**Figure 2:**
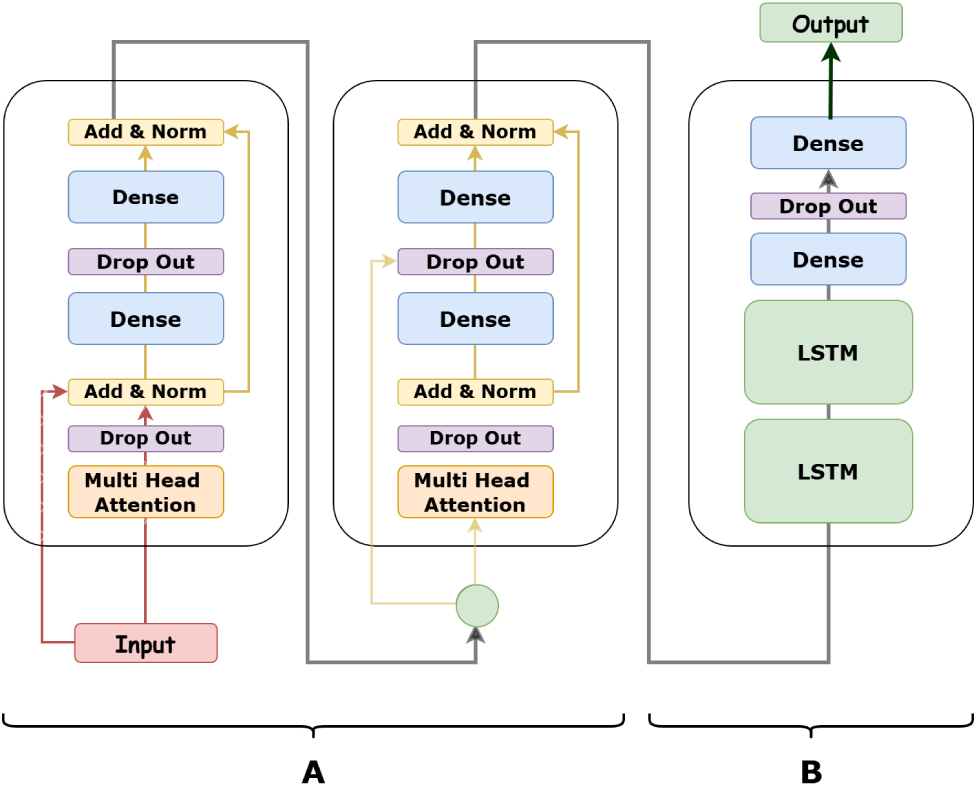
The proposed vmodel combines attention and memory to learn important patterns over time. This hybrid architecture captures both long-range dependencies and temporal progression in patient trajectories.Block A consists of stacked Transformer encoder layers with multi-head attention and residual normalization, while Block B integrates LSTM layers followed by dense projections to produce the final AKI prediction.

**Figure 3:**
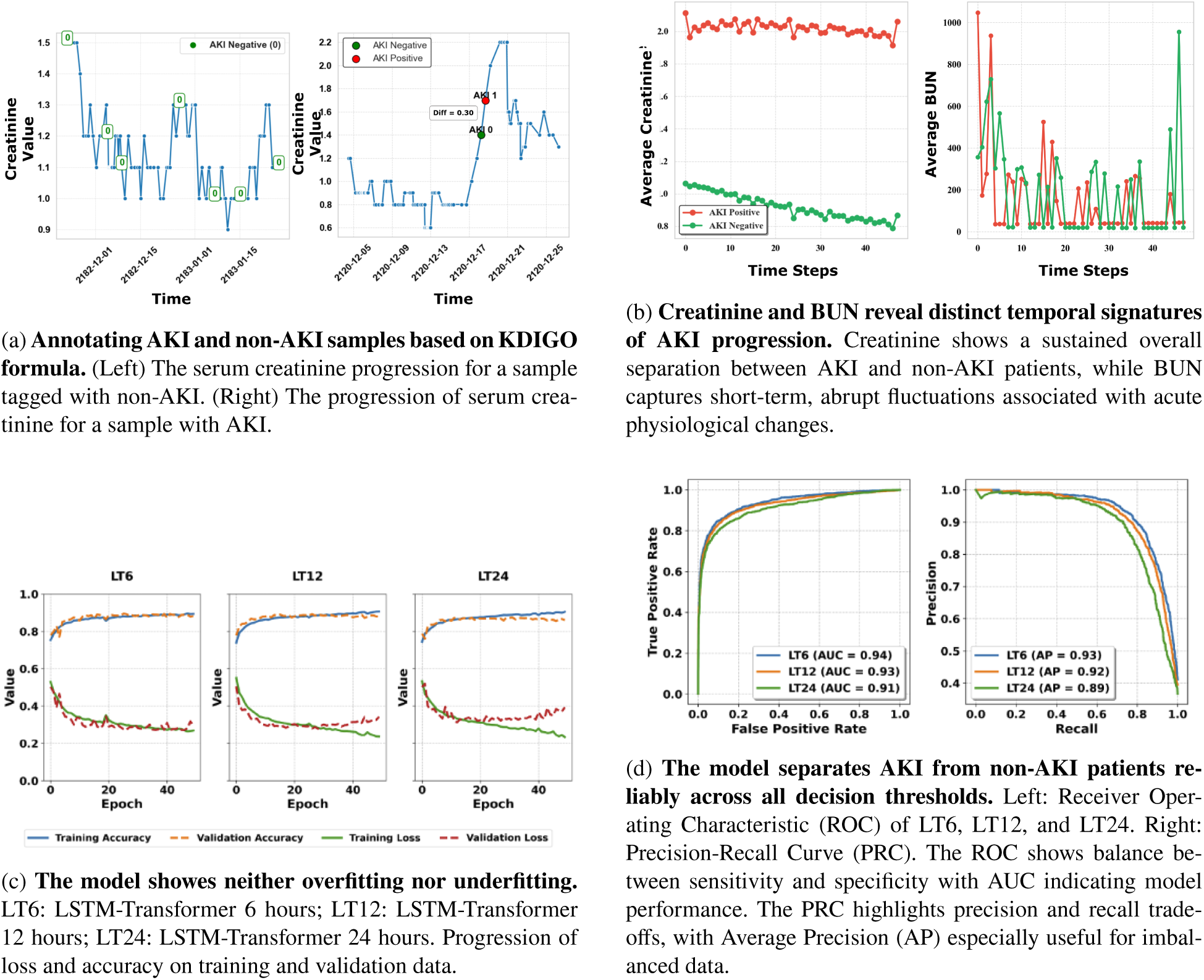
Model behavior and performance analysis. (A) AKI vs non-AKI labeling based on KDIGO criteria. (B) Temporal trends of creatinine and BUN. (C) Training and validation loss/accuracy showing stable learning. (D) ROC and PRC curves demonstrating strong predictive performance.

**Table 3.** Model performance is robust to training parameters. Even when the learning rate or batch size changes, most precision, recall, and F1 scores stay close to each other, showing the model works well in many settings.

| Time | 6 hrs |  |  | 12 hrs |  |  | 24 hrs |  |  |
| --- | --- | --- | --- | --- | --- | --- | --- | --- | --- |
|  | Prec. | Rec. | F1 | Prec. | Rec. | F1 | Prec. | Rec. | F1 |
| **Learning Rate** |  |  |  |  |  |  |  |  |  |
| 0.001 | **0.87** | **0.85** | **0.86** | 0.86 | 0.83 | **0.84** | **0.81** | 0.80 | 0.80 |
| 0.0001 | **0.87** | 0.82 | 0.84 | **0.93** | 0.77 | **0.84** | **0.81** | 0.80 | **0.81** |
| 0.00001 | 0.70 | 0.82 | 0.76 | 0.78 | 0.69 | 0.73 | 0.69 | 0.78 | 0.73 |
| **Batch Size** |  |  |  |  |  |  |  |  |  |
| 32 | 0.86 | 0.80 | 0.83 | 0.82 | 0.82 | 0.82 | 0.80 | **0.84** | **0.82** |
| 64 | **0.88** | 0.81 | 0.84 | **0.84** | **0.84** | **0.84** | **0.81** | 0.80 | 0.80 |
| 128 | 0.84 | **0.86** | **0.85** | 0.77 | 0.76 | 0.77 | 0.77 | 0.83 | 0.80 |

**Table 4.** Combining Transformer and LSTM yields consistently better and more balanced performance. The LSTM alone loses global context, leading to high recall but low precision, while the Transformer alone showes oposite result. The combination keeps both long-range and short-range information, giving a more balanced result

| Time | Only LSTM |  |  | Only Transformer |  |  | Transformer+LSTM |  |  |
| --- | --- | --- | --- | --- | --- | --- | --- | --- | --- |
|  | Prec. | Rec. | F1 | Prec. | Rec. | F1 | Prec. | Rec. | F1 |
| 6 hr | 0.68 | 0.89 | 0.77 | 0.89 | 0.79 | 0.84 | 0.88 | 0.81 | 0.84 |
| 12 hr | 0.66 | 0.62 | 0.64 | 0.92 | 0.76 | 0.84 | 0.84 | 0.84 | 0.84 |
| 24 hr | 0.64 | 0.52 | 0.57 | 0.82 | 0.78 | 0.80 | 0.81 | 0.80 | 0.82 |

**Table 5.** Top 12 Risk Factors for AKI. Both Integrated Gradients and counterfactual analysis consistently identify established clinical biomarkers, demonstrating that our model learns clinically valid predictive patterns.

| Risk | Integrated Gradients | Counterfactual Analysis |
| --- | --- | --- |
| Creatinine | 6.10e-02 | 0.034368 |
| BUN | 5.16e-03 | 0.000015 |
| Phosphate | 3.21e-03 | 0.007303 |
| RDW | 1.57e-03 | 0.002526 |
| Glucose | 1.55e-03 | 0.002041 |
| PT | 1.13e-03 | 0.000918 |
| INR | 1.37e-03 | 0.000750 |
| pCO<sub>2</sub> | 1.33e-03 | 0.000852 |
| Urea Nit. | 1.77e-03 | 0.000263 |
| Magnesium | 2.40e-03 | 0.000102 |
| Anion Gap | 1.14e-03 | 0.000029 |
| Potassium | 1.74e-03 | -0.000132 |

#### 4.6.2. Removing the LSTM

When the LSTM is removed, the model relies solely on the Transformer module (see Fig. 2 A). The performance of this setup is shown in Table 4. Using only the Transformer yields strong precision and F1-scores, particularly at the 6-hour and 12-hour horizons (F1 = 0.84 for both). However, recall is slightly lower than that of the full Transformer-LSTM model, especially for the 6-hour horizon. This suggests that while the Transformer is effective at extracting meaningful features, it lacks the sequential modeling capacity of the LSTM, which is more effective at capturing longer-term dependencies, particularly important for 24-hour prediction.

The ablation study shows that Transformer and LSTM contribute to the model’s performance. The Transformer captures short-term dependencies and provides strong feature extraction, while the LSTM excels at modeling long-term temporal dependencies. The combined Transformer-LSTM model outperforms both components, underscoring the importance of integrating these mechanisms for robust AKI prediction across different time horizons.

### 4.7. Modifiable Risk Factor Analysis (RQ4)

We analyze the model to identify modifiable risk factors that affect AKI prediction, enabling clinicians to intervene within the prediction horizon. Modifiable risk factors are defined as physiological laboratory or treatment-related variables that can be influenced through routine ICU management. Demographic and contextual variables such as age, gender, admission type, and hours since ICU admission are excluded because they cannot be modified during patient care.

To quantify the contribution of modifiable risk factors, we apply integrated gradients to the trained RenalTransLSTM model [30]. Integrated Gradients explains a model’s prediction by measuring how the predicted AKI risk changes as each input feature moves from its baseline value to the patient’s actual value. A baseline is defined as the median feature value in the training cohort for each time step. Time-resolved attributions are aggregated across the 48-hour observation window and across patients.

To further evaluate clinical actionability, we perform a counterfactual analysis [6] by simulating a modest reduction of ∼ 20% in each modifiable risk factor throughout the 48-hour window while keeping all other features unchanged. We then measure the average change in predicted AKI risk.

We identify the 25 most important risk factors for AKI using our interpretability methods. Among these, 12 (see Table.5) were consistently highlighted by both integrated gradients and counterfactual analysis, making them reliable predictors. Most of these are well-known clinical biomarkers. For example, serum creatinine and blood urea nitrogen (BUN) are widely used to monitor kidney function [16, 33]. Electrolytes like phosphate, potassium, and magnesium indicate kidney and metabolic health [34]. Glucose reflects metabolic stress, while coagulation measures such as PT and INR indicate overall physiological status [34, 36]. Red cell distribution width (RDW) is linked to inflammation and anemia [14], and blood gas measures like pCO_2_ show respiratory and acid–base balance [34]. The fact that our model identifies these known biomarkers suggests it is capturing meaningful clinical signals rather than random patterns. Other risk factors found by the model may also reveal new or complementary predictors of AKI, which could be explored in future studies.

## 5. Discussion

Our results show that the proposed RenalTransLSTM model can predict acute kidney injury at an early stage in ICU patients. Using 48 hours of patient history, the model can forecast AKI 6, 12, and 24 hours before it happens. The model achieved strong performance across all time horizons, with high AUC and balanced precision and recall even when the data is imbalanced.

These findings build on previous work in machine learning for AKI prediction. Earlier approaches such as logistic regression, random forest, and gradient boosting have shown reasonable performance, but they treat patient data as static features. This limits their ability to capture how a patient’s condition changes over time. More recent deep learning models, such as LSTM and transformer based approaches, try to use temporal information. However, many of these models focus on a single prediction time or do not fully combine short term and long term patterns.

Our approach addresses these limitations by combining LSTM and transformer in a single framework. The LSTM helps capture sequential changes in patient data, such as gradual increases in creatinine. The transformer helps capture relationships across the entire time window. Together, they provide a more complete understanding of patient state. This combination leads to better performance and more stable predictions across different time horizons.

Another important contribution of this work is the focus on clinically useful prediction windows. Instead of predicting AKI at the time of onset, the model predicts it several hours in advance. This allows clinicians to take action before the condition becomes severe. Early prediction can help reduce complications, lower the need for dialysis, and improve patient outcomes.

We also examined which risk factors are important for the model. The model consistently identified known clinical biomarkers such as creatinine, blood urea nitrogen, and electrolyte levels. These are widely used in clinical practice. The fact that the model highlights these factors suggests that it is learning meaningful clinical patterns rather than random signals. In addition, some other factors identified by the model may provide new insights and can be explored in future work.

There are some limitations in this study. The model was trained and tested on a single dataset, which may limit how well it works in other hospitals or populations. External validation using data from multiple centers is needed. In addition, the transformer component increases computational cost, which may make real time deployment more challenging in some settings. Although we improved interpretability using feature analysis, more work is needed to make the model outputs easier for clinicians to understand and trust.

In future work, we plan to include additional types of data such as clinical notes, imaging, and other patient records. We also aim to test the model on larger and more diverse datasets. Another direction is to integrate the model into a real time clinical system, where predictions can support decision making at the bedside.

In summary, RenalTransLSTM provides a strong framework for early AKI prediction. By combining temporal modeling with global context, it improves prediction accuracy and provides clinically useful early warnings. This approach has the potential to support better decision making in critical care and can be extended to other time sensitive clinical problems.

## Data Availability

The data used in this study were obtained from the publicly available MIMIC-IV database. Access to the dataset requires completion of the required training and a data use agreement.

https://physionet.org/content/mimiciv/3.1/

## 6. Acknowledgements

This work was supported by a College of Information Seed Grant at the University of North Texas.

